# IRE1α-XBP1 Activation Elicited by Viral Singled Stranded RNA via *TLR8* May Modulate Lung Cytokine Induction in SARS-CoV-2 Pneumonia

**DOI:** 10.1101/2022.01.26.22269752

**Authors:** José J. Fernández, Cristina Mancebo, Sonsoles Garcinuño, Gabriel March, Yolanda Alvarez, Sara Alonso, Luis Inglada, Jesús Blanco, Antonio Orduña, Olimpio Montero, Tito A. Sandoval, Juan R. Cubillos-Ruiz, Elena Bustamante, Nieves Fernández, Mariano Sánchez Crespo

**Author notes:** Corresponding author (M.S.C.). These authors contributed equally to this work (Nieves Fernández, Mariano Sánchez Crespo).

## Abstract

Initial symptoms of COVID-19 infection depend on viral replication, while hyperinflammation is a hallmark of critical illness and may drive severe pneumonia and death. Among the mechanisms potentially involved in the hyperinflammatory state, we focused on the unfolded protein response, because the IRE1α-XBP1 branch can be activated as result of the endoplasmic reticulum stress produced by the overwhelming synthesis of viral components and synergizes with Toll-like receptor signaling to induce cytokine expression. Viral RNA may trigger the IRE1α-XBP1 branch via TLR7/8 activation and like TLR2 and TLR4 may underpin cytokine expression trough *XBP1* splicing (*sXBP1*). The expression of *IL1B*, *IL6*, and *TNF* mRNA in bronchoalveolar aspirates (BAAs) were higher in COVID-19 patients under mechanical ventilation and intubation who showed *sXBP1*. The scrutiny of monocytic/macrophagic markers during active infection showed a reduction of those involved in antigen presentation and survival, as well as the IFN stimulated gene *MX1*. These changes reverted after infection tests turned negative. In contrast, the expression of the mRNA of the serine protease *TMPRSS2* involved in S protein priming showed a high expression during active infection. *TLR8* mRNA showed an overwhelming expression as compared to *TLR7* mRNA, which suggests the presence of monocyte-derived dendritic cells (MDDCs). *In vitro* experiments in MDDCs activated with ssRNA40, a positive-sense, single-stranded RNA (+ssRNA) like SARS-CoV-2 RNA, induced *sXBP1* and the expression of IL-1β, IL-6, and *TNF*α at mRNA and protein levels. These responses were blunted by the IRE1α ribonuclease inhibitor MKC8866. Given the analogies between the results observed in BAAs and the effects induced by +ssRNA in MDDCs, IRE1α ribonuclease inhibition might be a druggable target in severe COVID-19 disease.

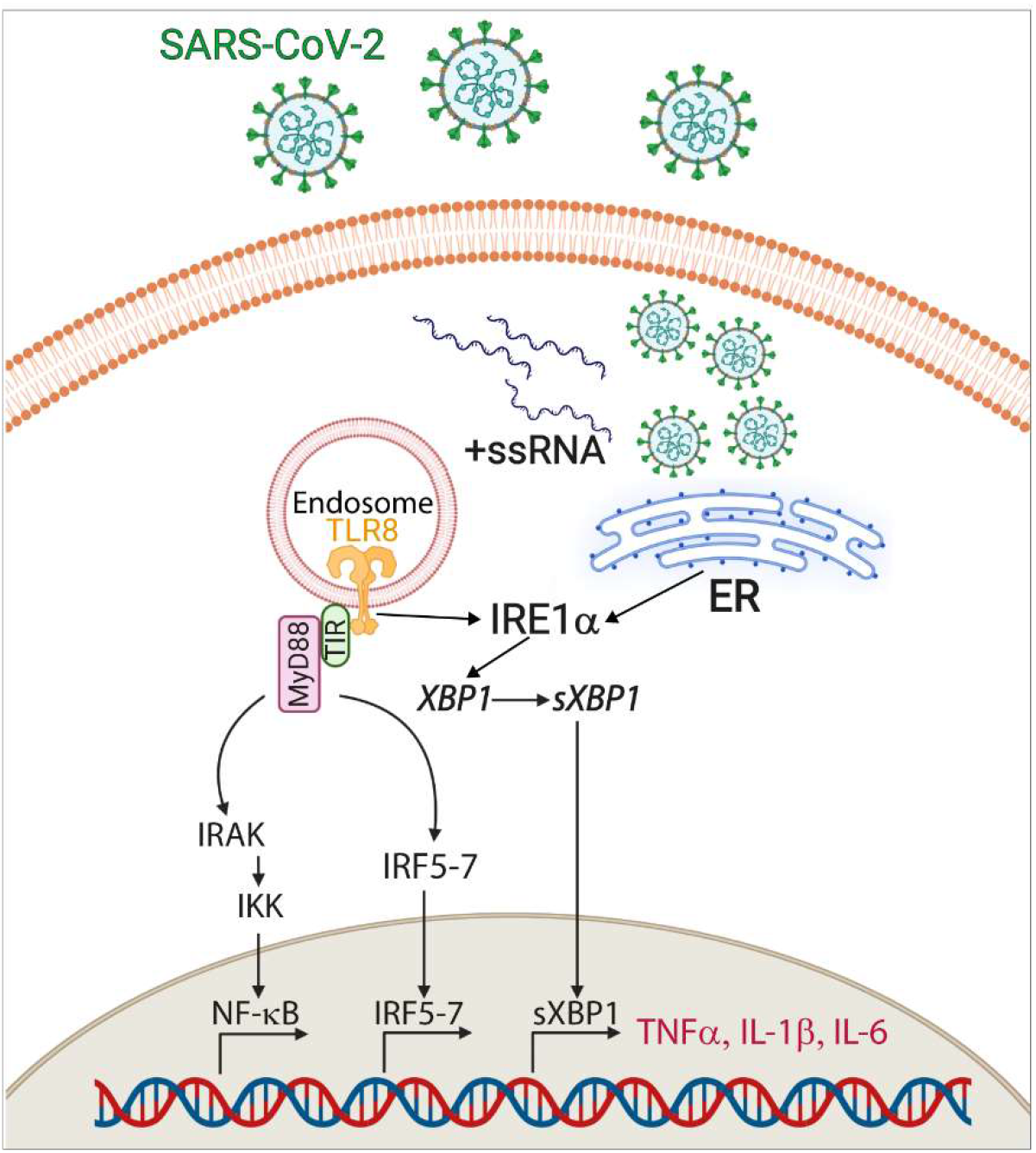

**Author summary:** COVID-19 pandemics put an unprecedented pressure on health systems. The need of new therapies urged research on the mechanisms triggered by the interaction of SARS-CoV-2 virus with host cells and the ensuing pathophysiology driving pneumonia and multiorgan failure. Hyperinflammation soon appeared as a mechanism involved in mortality that could even proceed after viral infection comes to an end. Hyperinflammation is supported by an inappropriate production of cytokines, and this explains the use of the term cytokine storm to refer to this phase of the disease. Given that insight into the molecular mechanisms driving cytokine storm should focus on the interaction of viral components with immune cells, experiments addressing the effect of viral components on its cognate receptors were carried out. It was observed that viral RNA induces a cytokine pattern like the one observed in bronchoalveolar aspirates of COVID-19 patients with critical disease. Overall, the study revealed that both cell organelle overload and receptors involved in the recognition of viral RNA may team up to induce proinflammatory cytokines. This mechanism can be exploited to develop new treatments for COVID-19 disease.

## Introduction

Coronavirus disease 2019 (COVID-19) is a pandemic infection produced by severe acute respiratory syndrome coronavirus 2 (SARS-CoV-2). Habitual evolution includes an initial influenza-like phase where fever and unproductive cough are predominant, but it can lead to severe respiratory insufficiency and multiorgan failure, which in many cases proceeds after infection resolution due to a hyperinflammatory response associated with the production of proinflammatory cytokines. The term cytokine storm was coined to refer to this condition [1], but the analysis of the immunopathological damage and the analogies with other viral diseases make it most appropriate the use of viral sepsis to depict the association of T-cell deficiencies with systemic hyperinflammation driven by virus-host cell interaction [2]. The use by viruses of the translational machinery of the host to produce their materials overloads many cellular functions, including protein folding and secretion in the endoplasmic reticulum (ER). Since only properly folded proteins should exit from the ER to maintain homeostasis, cells arrange a response directed to retain and degrade defective proteins. This involves an intracellular signaling pathway termed the unfolded protein response (UPR) [3–5]. The UPR includes a down regulation of global protein synthesis, the degradation of some proteins, and the transcriptional induction of specific genes associated with the activity of its three branches. The most conserved one depends on the dual activity of inositol-requiring enzyme 1α (IRE1α), which shows protein kinase and endonuclease activities [6]. The endonuclease activity cleaves a 26-base fragment of the mRNA of the preformed transcript of *XBP1*, which is followed by ligation of the resulting exons to yield *sXBP1*. This drives the translation of the functionally active factor *sXBP1* that *trans*-activates genes encoding proinflammatory proteins, e.g., the prostaglandin producing enzymes cyclooxygenase 2 and the microsomal isoform of prostaglandin synthase 2 [7], the ubiquitin-like modifier ISG15, and the cytokines IFNβ, IL-6, IL-23, and *TNF*α [8–11].

Although the goal of the UPR is alleviating ER-stress, the IRE1α-*XBP1* branch contributes to the pathogenesis of multiple ailments, including cancer, atherosclerosis, infections, and autoimmune diseases [12–15]. Recent research has disclosed that the IRE1α-*XBP1* branch enhances viral transcription and contributes to maintain dormant viral genomes in a host of infections [16, 17]. The notion that UPR branches can mediate immune recognition of zoonotic viruses and drive acute lung injury through ER-stress mediated inflammation was disclosed by Hrincius et al. [18], who showed activation of the UPR by infection with poorly glycosylated pandemic strains of influenza A virus. The IRE1α route was recruited, and its activation reduced when glycans were added to specific sites in the globular head of hemagglutinin. A study in cell lines infected with a SARS-CoV-2 isolate and lung biopsies of COVID-19 patients showed that SARS-CoV-2 hijacks the glycosylation biosynthetic machinery and drives ER-stress and UPR activation. This is due to the overwhelming asparagine N-linked glycosylation required for the translation of viral glycoproteins, which forces aberrant glycosylation and ER-stress [19]. In keeping with notion, specific activation of *sXBP1* has been reported in monocytes from COVID-19 patients [20].

The association of *sXBP1* with TLR2 and TLR4 signaling [8] together with genuine UPR induced by protein synthesis overload point to *sXBP1* involvement in the cytokine storm. [21]. The role of cytokine storm in COVID-19 disease was supported by the beneficial effect of tocilizumab, a humanized antihuman IL-6 receptor antibody [22]. This is noteworthy given the dependence of IL-6 production on the IRE1α-*XBP1* branch [8] and the presence in the proximal promoter of *IL6* of at least six sequences binding to *sXBP1* [23]. Another potential interference of *sXBP1* on COVID-19 illness stems from its blunting effect on dendritic cell homeostasis and the metabolic fitness of T-cells [24, 25]. Likewise, the σ1 receptor (S1R) reduces cytokine production in murine models of septic shock through IRE1α endonuclease activity inhibition [14]. A new piece of evidence associating the UPR with COVID-19 illness was the induction of the UPR by ORF8 and S viral proteins, as well as the reduction of viral replication by pharmacological inhibition of the IRE1α and ATF6 branches in epithelial cell lines [26]. In contrast, up-regulation of IRE1α (RNase) activity by cannabidiol has been found to block SARS-CoV-2 replication in a cell line derived from lung epithelial cells and in lungs and nasal turbinates of infected mice [27].

Given that SARS-CoV-2 is a +ssRNA virus, the tandem *TLR7*/8 comes into prominence given its ability to bind viral RNA. Consistent with this notion, loss-of-function variants in X-chromosomal *TLR7* have been reported in young patients with severe COVID-19 disease, who showed impaired type I and II IFN responses [28]. The purpose of this study has been addressing how *sXBP1* and *TLR7*/8 engagement may underpin viral sepsis in COVID-19 disease. To this end, samples of nasopharyngeal swabs and bronchoalveolar aspirates (BAAs) were studied to address the presence of *sXBP1*, the cytokine-signature, and the expression of monocytic lineage cell markers and enzymes involved in energetic metabolism. After obtaining a profile of the transcriptional landscape, *in vitro* experiments were performed to address the transcriptional and energetic patterns of MDDCs stimulated with *TLR7* and *TLR8* agonists. Experiments showed that *TLR8* activation of MDDCs induces a pattern of *XBP1* splicing and cytokine expression, sensitive to inhibition of IRE1α RNase activity, which mimics the pattern observed in BAAs.

## Results

### Studies in Nasopharyngeal Samples

Initial assays were conducted in nasopharyngeal samples from patients receiving medical assistance for symptoms consistent with COVID-19 disease (Fig 1A). The extracted RNA was used for the diagnosis of SARS-CoV-2 infection and residual samples used for the assay of *sXBP1* by RT-PCR assays. This entails the separation of the PCR products by electrophoresis in agarose gel and densitometric analysis of GelRed stained bands. *sXBP1* is distinguished from unspliced *XBP1* (*uXBP1*) by its faster migration. The position of the primers and the spliced region are shown in Fig 1B. These correspond to GenBank sequence NM_001079539.2, which differs from *uXBP1* sequence NM_005080.4 by the deletion of 26 nucleotides. Separate sequencing of the bands was used to confirm splicing (Fig 1C). A random selected array of RT-PCR negative and positive samples shows the presence of *uXBP1* and *sXBP1* (Fig 1D). The presence of three bands in some cases is explained by the formation of heteroduplexes [29]. *sXBP1* was detected in 17.91% of SARS-CoV-2 negative and 40.32% of SARS-CoV-2 positive patients (Fig 1E). Quantitation of *sXBP1* showed higher values in COVID-19 positive patients as compared to the negative ones (Fig 1F). The incidence of *sXBP1* was similar in male and females (Fig 1G) and increased with age (Fig 1H). Mortality was observed in four patients who showed a degree of splicing above 10% of total *XBP1* (Fig 1I). These findings show that *sXBP1* shows higher frequency and extent in nasopharyngeal exudates of patients with active SARS-CoV-2 infection, particularly in dying patients.

**Fig 1.**
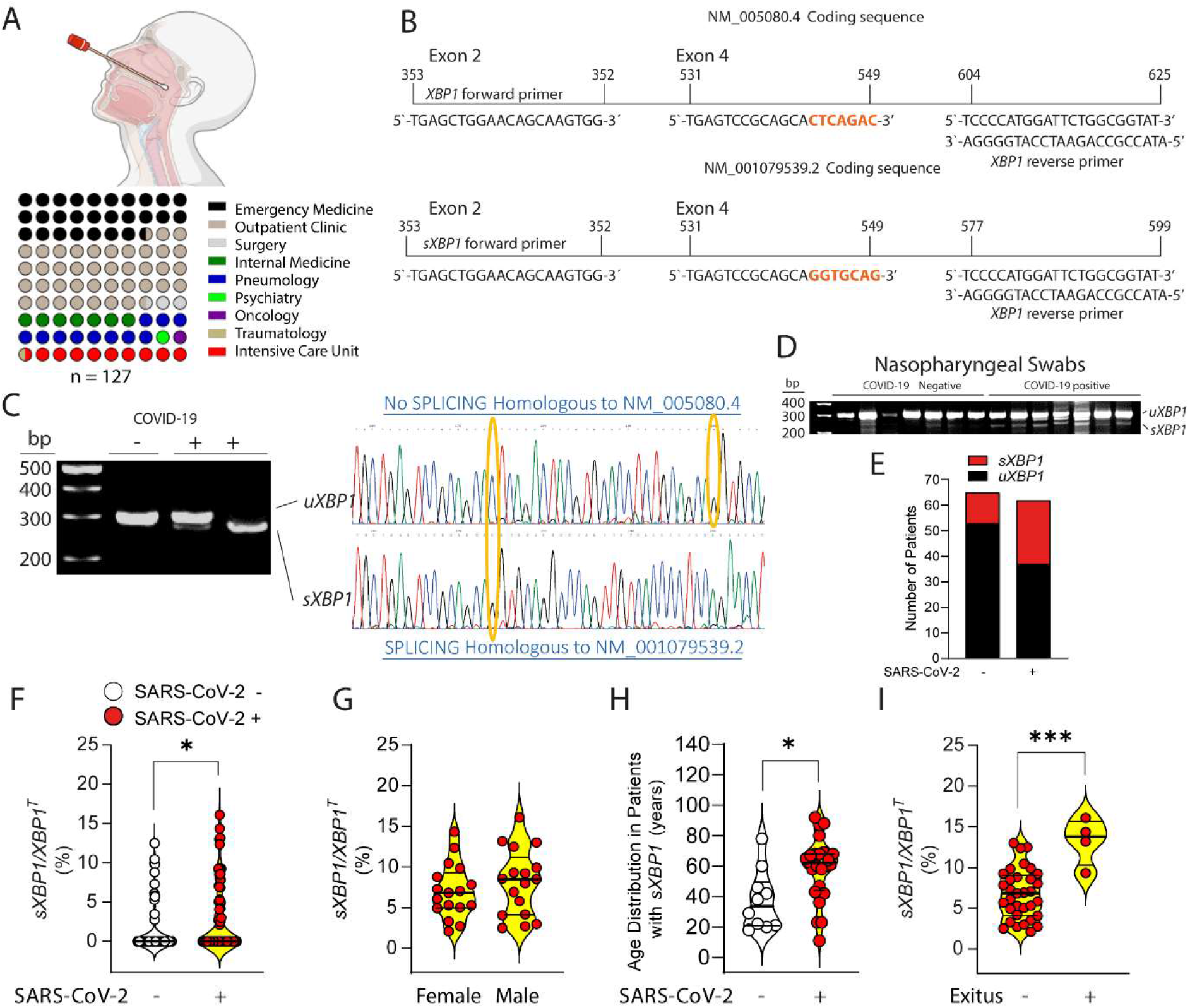
Sequences of *XBP1* mRNA transcripts and analysis of RT-PCR products in nasopharyngeal samples. (A) Medical departments involved in the obtention of nasopharyngeal swabs. (B) The splicing of 26 nucleotides in NM_005080.4 sequence generates the 531-549 sequence in NM_001079539.2. The position of primers, including the reverse primer spanning the spliced sequence is shown. (C) Agarose gel electrophoresis and sequencing of the amplicons obtained by RT-PCR with *XBP1* primers showing the migration of the spliced (*sXBP1*) and unspliced products (u*XBP1*). (D) The electrophoretic pattern of *XBP1* in a series of samples from COVID-19 negative and positive patients is shown. The presence of three bands in some cases is due to the formation of heteroduplexes. (E) Distribution of patients according to the presence or absence of *sXBP1* in SARS-CoV-2 negative and positive patients. (F) Quantitation of *sXBP1* versus total *XBP1* (*XBP1*^T^) in COVID-19 positive and negative samples. (G) Quantitation of *sXBP1* versus total *XBP1* in male and female SARS-CoV-2 positive patients. (H) Age distribution of patients with *sXBP1*(I) Quantitation of *sXBP1* versus total *XBP1* in SARS-CoV-2 positive according to the outcome. *p < 0.05, ***p<0.005, paired or unpaired (two-tail) t test.

### *sXBP1* and Cytokine Expression in BAAs

Further experiments were carried out using RNA extracted from BAAs of patients under mechanical ventilation in intensive care unit (ICU) (Fig. 2A). Ventilatory support and endotracheal intubation were indicated because of acute hypoxemic respiratory failure despite high-flow nasal oxygen therapy or non-invasive ventilation. COVID-19 patients received a standard and proved useful treatment for the hyperinflammatory state consisting of 6 mg dexamethasone daily or 50 mg of IV hydrocortisone every 8 hours for up to 10 days, while this protocol was not routinely used in non-COVID-19 patients (Fig 2B). The extent of *sXBP1* was higher in SARS2-CoV-2 pneumonia patients than in those with respiratory failure due to other conditions, decreased after COVID-19 tests turned negative, and showed higher values than those observed in nasopharyngeal swabs (Fig 2C). The PERK-eIF2α-ATF4-CHOP branch of the UPR was explored assaying *DDIT3*/CHOP gene expression. *DDIT3* expression also decreased after SARS-CoV-2 tests turned negative, while there was no significant difference of expression between non-COVID-19 and COVID-19 pneumonia patients (Fig 2D). As regards cytokine expression (Fig 2E-2N), *IL1B* and *IL6* mRNA levels during viral proliferation were significantly lower than those detected in non-COVID-19 patients. A similar trend was observed in *TNF*, *IL23A*, and *IL8* mRNA expression, although these values did not reach statistical significance. Cytokine mRNA did not show a trend to decrease after SARS-CoV-2 tests were negative. *IL10* and *IFNB* mRNA were higher in SARS-CoV-2 infection than in non-COVID pneumonia and continued elevated after COVID-19 tests turned negative. *IFNG* showed a trend to be increased in COVID-19 pneumonia. Overall, these results show that *sXBP1* in respiratory samples from COVID-19 patients does not associate with levels of cytokine expression higher than those observed in non-COVID-19 pneumonias. The high expression of *IL10* mRNA suggests a parallel activation of an archetypal anti-inflammatory cytokine that might counter the inflammatory response. The increased expression of *IFNB1* mRNA is consistent with its involvement in viral sepsis.

**Fig 2.**
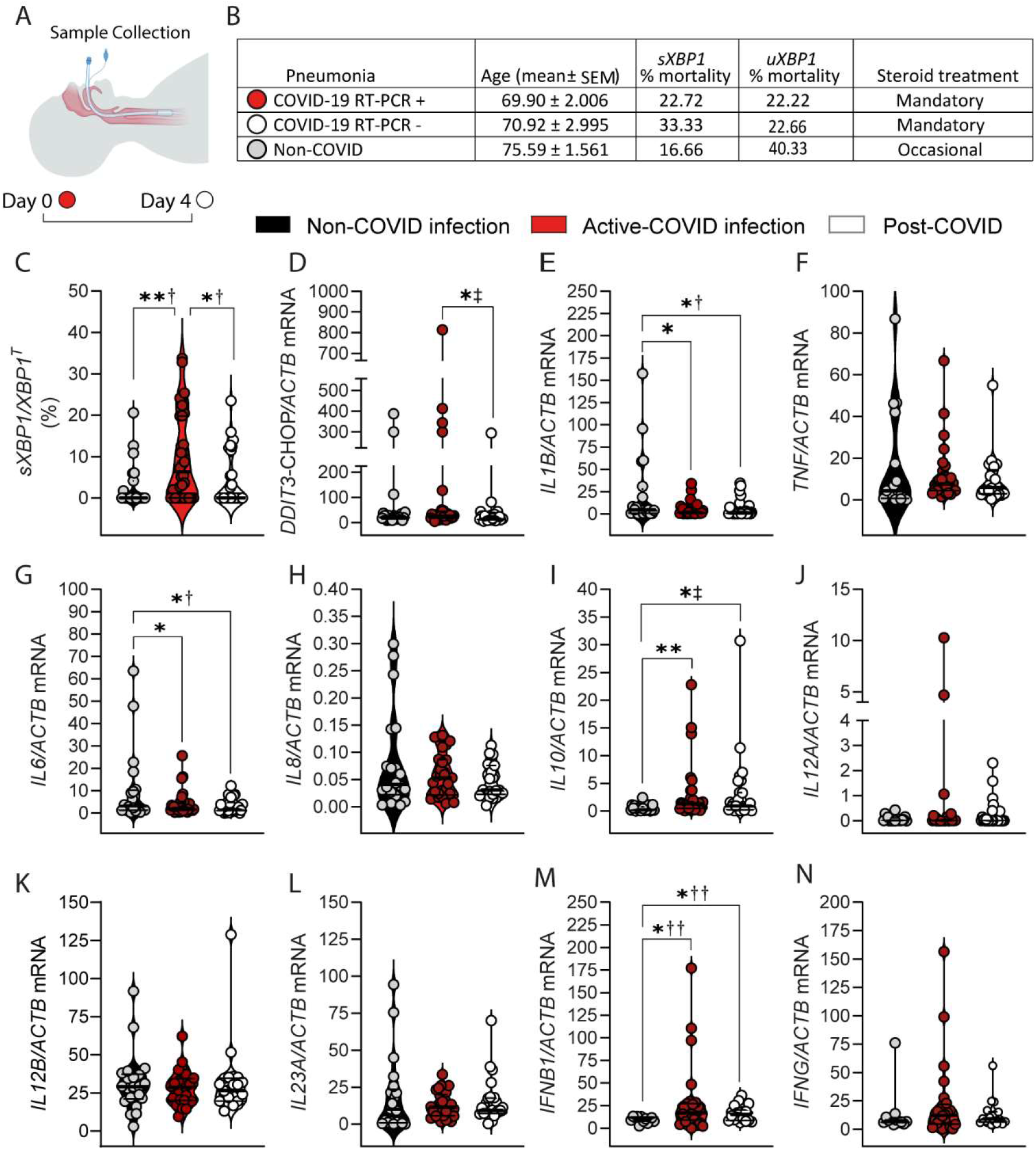
Expression of the mRNA of *XBP1*, *DDIT3*/CHOP, and cytokines in BAAs of patients under mechanical ventilation in ICU. (A) Scheme of sample collection. (B) Stratification of patients according to non-COVID-19, active COVID-19, and non-active COVID-19 infection. (C) *sXBP1* expression in the different cohorts. Results are expressed as mean ± SEM. †Ordinary one-way ANOVA with the Tukey’s multiple comparisons test. *p < 0.05. *** p < 0.005. (D) Expression of the mRNA de *DDIT3*/CHOP. ‡Kruskal-Wallis *U* test. (E-N) Expression of the mRNA encoding *IL1B*, *TNF*, *IL6*, *IL8*, *IL10*, *IL12A*, *IL12B*, *IL23A*, *IFNB1*, and *IFNG* in BAAs. Data are presented as mean ± SEM. *p < 0.05, **p < 0.01, ***p<0.005. †Ordinary one-way ANOVA. ‡Kruskal-Wallis *U* test. ††Welch and Brown-Forsythe ANOVA test.

### *sXBP1* Stratification in SARS-CoV-2 Patients during ICU Hospitalization

SARS-CoV-2 patients were stratified according to the presence or absence of both active infection and *sXBP1*Fig 3A shows viral load in samples collected during infection and at the time when a negative COVID-19 test was first recorded. *PTGS2* mRNA was higher in patients with *sXBP1* both during infection and after negativization of the RT-PCR test (Fig 3B). *TNF* and *IL1B* mRNA expression was also higher in patients with *sXBP1*, even after a negative RT-PCR test (Fig 3C and 3D). *IL6* mRNA was increased in patients with *sXBP1* and active infection (Fig 3E). IL8 mRNA was expressed at much lower levels than those encoding other cytokines, particularly after negativization of the infection in patients who did not show *sXBP1* (Fig 3F). *IL10* mRNA was higher in SARS-CoV-2 positive patients who did not show *sXBP1* and remained elevated in patients showing *sXBP1* after resolution of the infection. These results show a high expression of the mRNA of *PTGS2* and several proinflammatory cytokines in patients with *sXBP1*, which may persist after SARS-CoV-2 test becomes negative. The sustained expression of *PTGS2* (COX2) mRNA suggests its involvement in either induction or repair of inflammatory damage [30]. Together, the results agree with the reported role of *sXBP1* in the transcriptional activation of COX2, *TNF*α, IL-1β, and IL-6.

**Fig 3.**
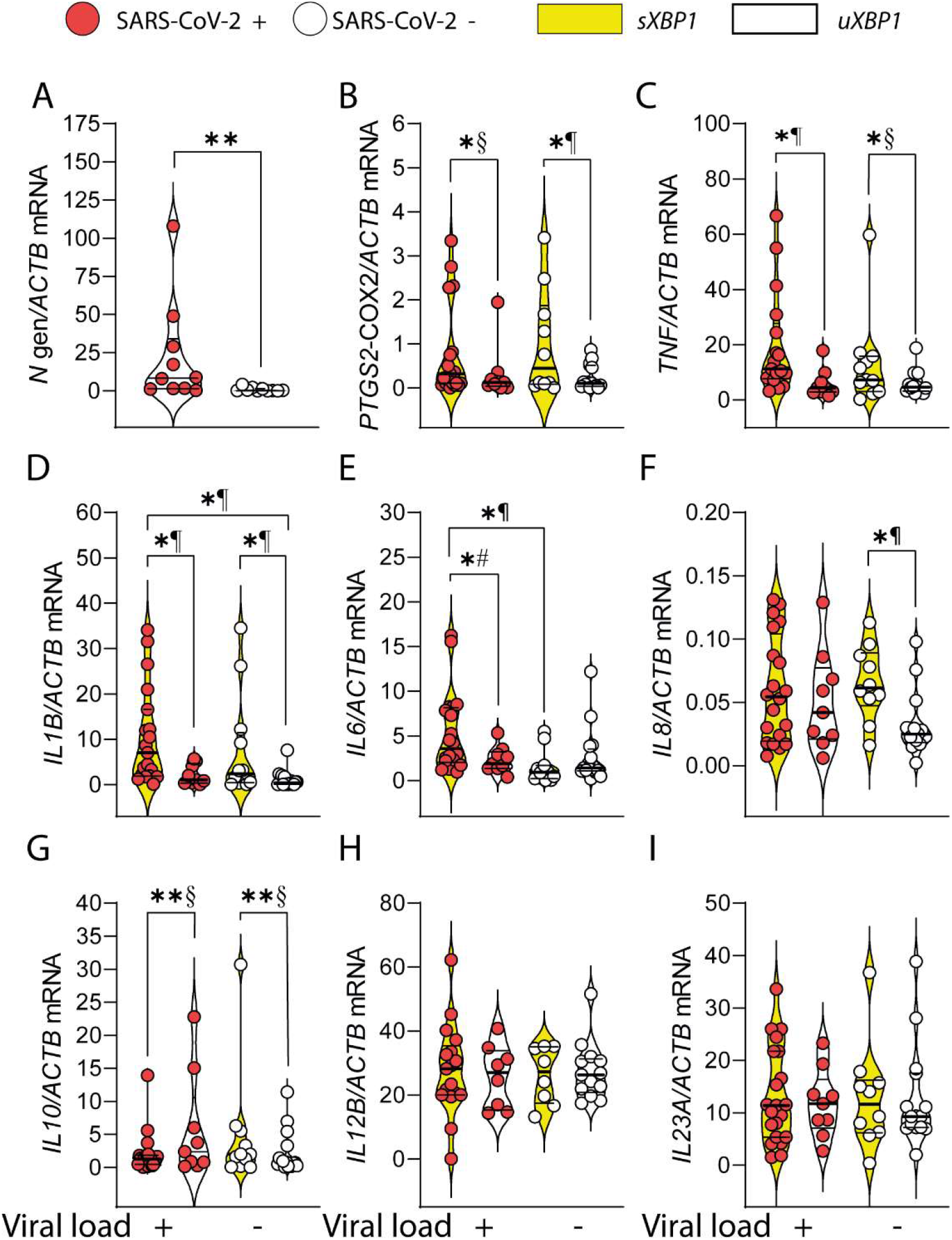
Association of *sXBP1* splicing and viral replication with cytokine expression in BAAs. (A) Viral load in samples obtained at the time of SARS-CoV-2 positive and negative tests. Data are presented as mean ± SEM. **p < 0.01. Unpaired two-tailed Student’s *t* test. (B-I) Patients were stratified in cohorts according to the presence of *sXBP1* and the presence or absence of viral load as deemed from RT-PCR test for SARS-CoV-2 infection. The mRNA of *PTGS2* and various cytokines was assayed in the extracted RNA and the statistical significance of the results was assayed using ordinary one-way ANOVA with Tukeýs post-hoc multiple comparison test. Data are presented as mean ± SEM. *p<0.05, **p<0.01, ***p<0.005. §One-sample Wilkoxon signed rank test. ¶Unpaired (two-tail) *t* test. #Welch’s test.

Expression of Genes Involved in Glycolysis and Oxidative Phosphorylation (OXPHOS) Lymphocytes and myeloid cells respond to pathogen-associated molecular patterns (PAMPs) with a robust rewiring of their energetic metabolism driving increased glycolysis (Fig 4A). The impairment of O_2_ supply due to pneumonia further explains the resort to glycolysis and agrees with reports showing that SARS-CoV-2-induced metabolic reprogramming enhances the production of proinflammatory cytokines and IFNs by monocytes, and concomitantly inhibits T cell function [31, 32]. Consistent with this notion, the expression of *GLUT1* mRNA, a glucose transporter, and *HIF1A* mRNA, a transcription factor involved in the regulation of glycolytic enzymes, were increased during active infection. However, there was no difference as compared to non-COVID-19 pneumonia (Fig 4B and 4C). The mRNA encoding hexokinase II (Fig 4D), pyruvate dehydrogenase kinase IV (Fig 4F), malate dehydrogenase (MD) 2 (Fig 4G), and cis-aconitate dehydrogenase (*IRG1* gene) (Fig 4K) increased during active infection as compared to both non-COVID-19 pneumonia and post-COVID infection. Notably, proteins involved in mitochondrial function also increased during COVID-19 pneumonia, i.e., succinate dehydrogenase subunit A (Fig 4I) and the 2-oxoglutarate-malate transporter SLC25A11, (Fig 4J). Together, these data show a resort to glycolysis during active SARS-CoV-2 infection that seems supported by the activity of HIF1and elements of the malate-aspartate shuttle such as MD2 and SLC25A11, which buttress the NAD^+^/NADH redox balance necessary for the progression of glycolysis at the glyceraldehyde 3-phosphate-dehydrogenase step.

**Fig 4.**
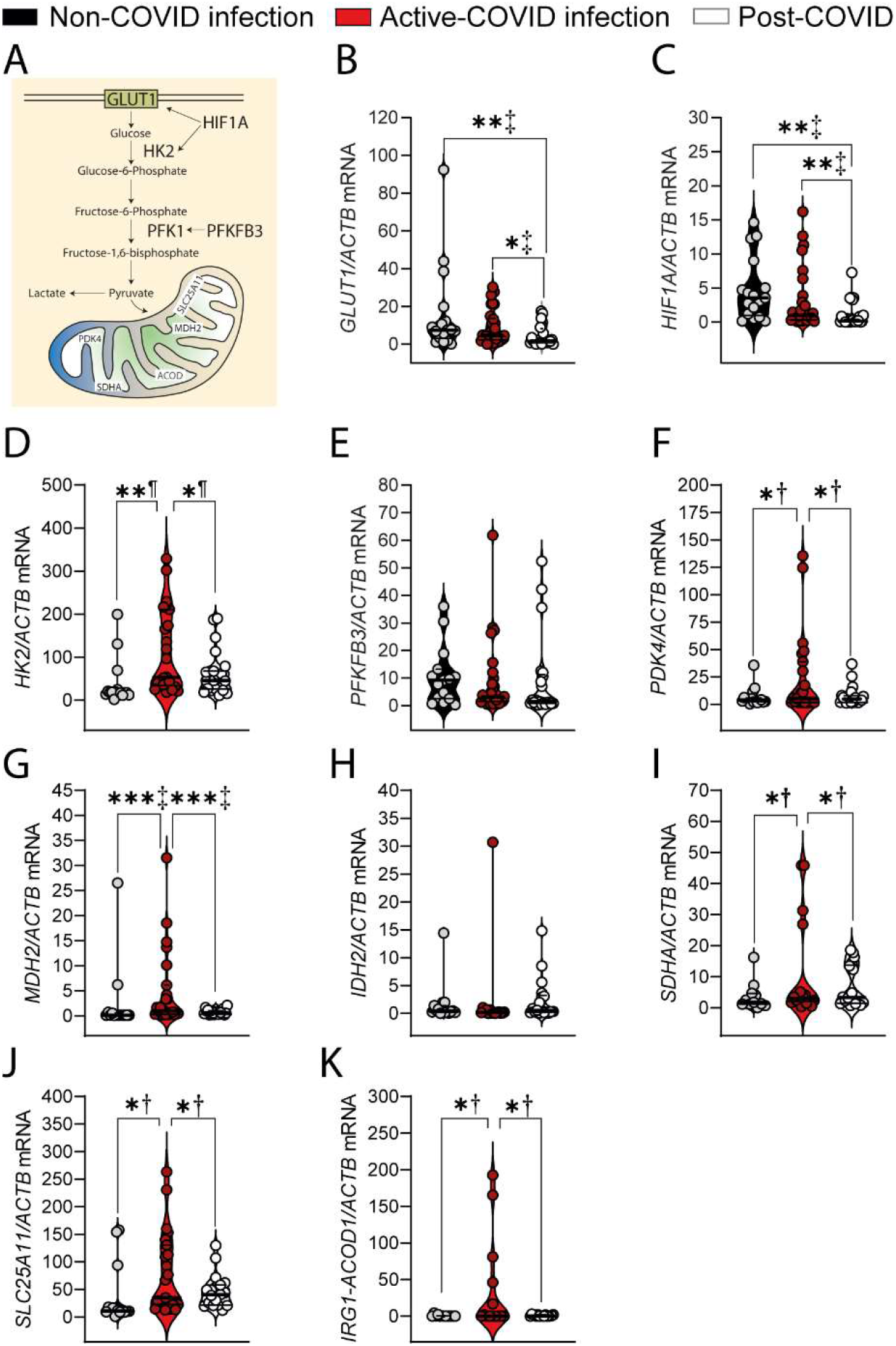
Expression of enzymes involved in glycolysis and mitochondrial proteins. (A) Diagram of glycolytic and mitochondrial proteins assayed in BAAs. (B-K) BAAs of patients with controlled respiration were used for RNA extraction and RT-PCR assay of mRNA expression of genes encoding for proteins involved in glycolysis, response to hypoxia, and mitochondrial function. *GLUT1*, glucose transporter 1. *HIF1A*, hypoxia-inducible factor 1α. *HK2*, hexokinase 2. *PFKB3*, 6-phosphofructo-2-kinase/fructose-2,6-biphosphatase. *PDK4*, pyruvate dehydrogenase kinase. *MDH2*, malate dehydrogenase 2. *IDH2*, isocitrate dehydrogenase 2. *SDHA*, succinate dehydrogenase protein subunit A. *SLC25A11*, mitocondrial 2-oxoglutarate-malate carrier. *IRG1*-ACOD1, immunoresponsive gene 1-aconitate decarboxylase. Data are presented as mean ± SEM. *p < 0.05, **p < 0.01, ***p<0.005. ‡Kruskal-Wallis *U* test. †Ordinary one-way ANOVA. ¶Paired or unpaired (two-tail) *t* test.

### Characterization of Monocytic Markers in BAAs

The first attempt to characterize the monocytic/macrophagic populations in BAAs focused on the expression of *TLR7*/8, given their involvement in the recognition of viral RNA. *TLR8* mRNA was expressed to a far greater extent than *TLR7* mRNA (Fig 5A), which agrees with the decay of *TLR7* expression during the differentiation to MDDCs [33]. Further assay of markers showed a diminished expression of *HLA-DRB1* (Fig 5B), a gene involved in antigen presentation, *CD300E* (Fig 5C), a gene associated with survival signals, the chemokine receptor *CCR2* (Fig 5D), and the IFN-stimulated gene (ISG) *MX1*, (Fig 5G). Negativization of viral replication was associated with an increase of the expression of these genes, as well as the migration receptor *MMP9* (Fig 5E) and the MDDC differentiation marker *BATF3* (Fig 5F). *PTGS2*/COX2 mRNA was similar in non-COVID and active COVID infection and decreased after RT-PCR test became negative (Fig 5I). The mRNA of the ISG *OAS1* did not show significant changes during SARS-CoV-2 infection and increased after negativization of SARS-CoV-2 test (Fig 5H). *TMPRSS2* mRNA, a serine protease involved in the cleavage of viral spike proteins [34] was significantly increased during SARS-CoV-2 active infection and (Fig 5J). These results disclose a differentiation profile during COVID-19 infection characterized by a low expression of markers associated with antigen presentation and survival signals, as well as *MX1*. This is followed by an increase of the markers expressed in MDDCs in response to TLR ligands [33]. The high expression of *TMPRSS2* mRNA underscores the role of *TMPRSS2* in SARS-CoV-2 cell invasion and the low expression of *MX1* agrees with the reported association of single nucleotide polymorphisms within *TMPRSS2* and near *MX1* gene with severe COVID-19 disease [35]. A cogent explanation for the low *MX1* expression could be an evasive strategy of SARS-CoV-2 to avoid and/or shut down type I IFN responses [36].

**Fig 5.**
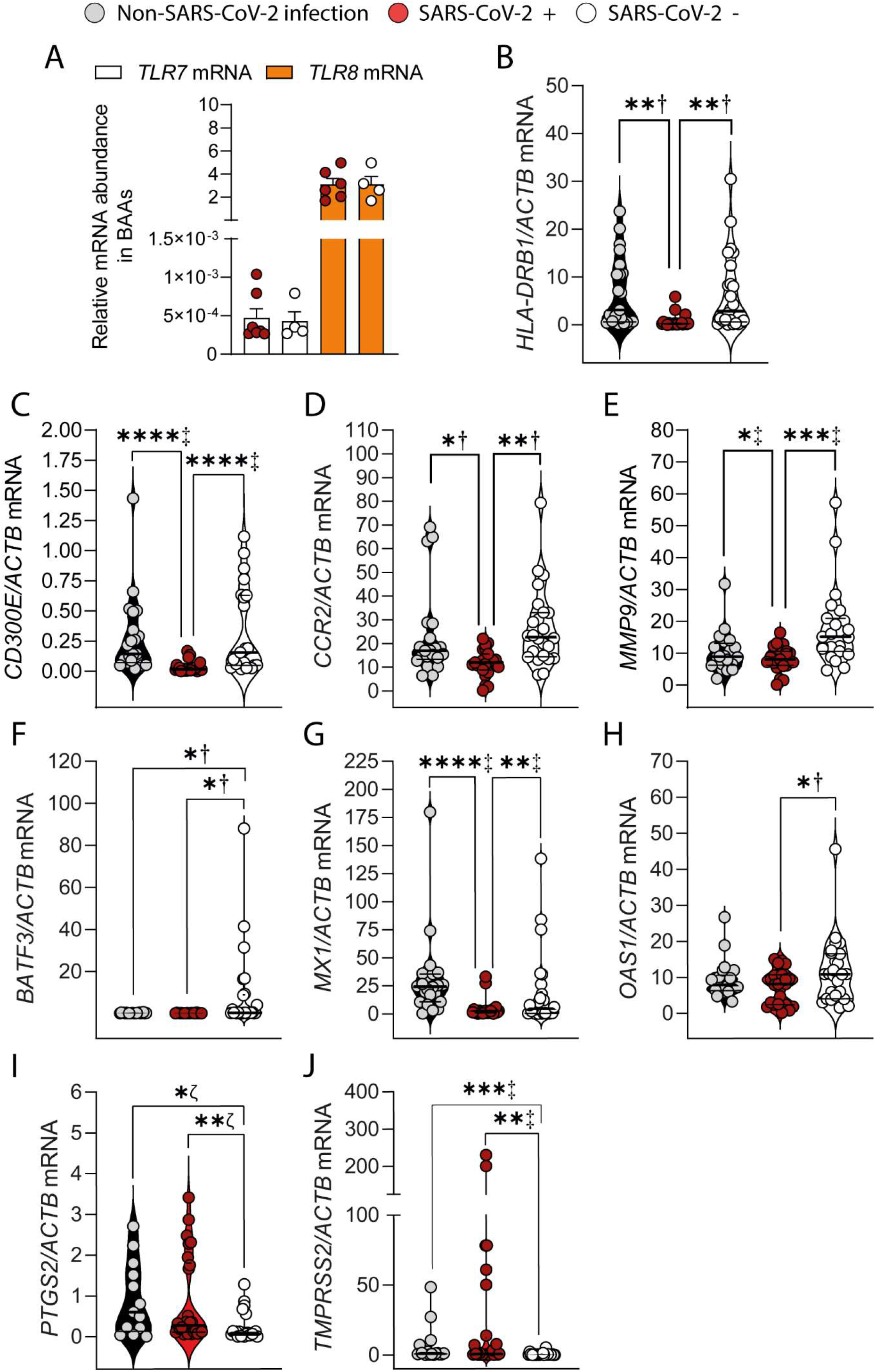
Expression of genes involved in monocytic-lineage differentiation in BAAs. (A) Expression of the mRNA encoding the receptors *TLR7* and *TLR8* in patients with active SARS-CoV-2 infection and after negative tests. (B-F) Expression of monocyte-differentiation markers in BAAs of non-COVID-19, COVID-19, and post-COVID-19 infection. (G-H) Expression of the ISG *MX1* and *OAS1* mRNA. (I) Expression of the mRNA encoding *PTGS2*/COX2. (J) Expression of the mRNA encoding the transmembrane serine protease *TMPRSS2*. Data are presented as mean ± SEM. *p < 0.05, **p < 0.01, ***p<0.005. †Ordinary one-way ANOVA. ‡Kruskal-Wallis U test. ζHolm-Sidak’s multiple comparison test.

### Effects Induced by the Stimulation of Receptors Involved in the Recognition of Viral RNAs in MDDCs

Because SARS-CoV-2 is a +ssRNA virus, we posited that *TLR7*/8 might shape the innate immune response, given their endosomal location and accessibility to intracellular viral RNA. *TLR7* and *TLR8* expression in MDDCs mimicked the pattern observed in BAAs by showing a high expression of *TLR8* and a low expression of *TLR7* mRNA (Fig 6A). Activation of *TLR7* by the selective ligand imiquimod showed a low extent of *XBP1* splicing, even in real-time RT-PCR assays using a reverse primer overlaping the spliced region (Fig 6B). Consistent with the effect of palmitate as a potentiator of imiquimod effect via metabolic rewiring and *XBP1* splicing [11], the expression of proinflammatory cytokines increased in the presence of palmitate (Fig 6C-6F). In contrast, 2-deoxyglucose, which enhances *XBP1* splicing in the presence of some PAMPs [10] showed a limited effect. Given that IFNγ teams up with *TNF*α to induce mortality in mice during SARS-CoV-2 infection [37] and it has been associated with the development of cytokine storm [38–41], the expression of ISGs was assayed. In contrast to IFNs, *MX1* and *OAS1* showed high levels of expression. The IRE1α RNase inhibitor MKC8866 [42] and the S1R agonist fluvoxamine, which has been reported to inhibit *XBP1* splicing in bacterial sepsis [14], lacked any significant effect on those responses (Fig 6G-6J). Real-time assays of energetic metabolism with the Seahorse technology, showed a reduction of O_2_ consumption rate (OCR) and an increased extracellular acidification rate (ECAR) in response to imiquimod (Fig 6K), thus mimicking the glycolytic rewiring induced by bacterial PAMPs [43]. These effects were enhanced by metformin, a well-known inhibitor of the complex I of the electron transport chain (Fig 6L) that has been associated with a beneficial effect on the evolution of COVID-19 disease [44], most likely explained by its ability to reduce IL-1β and enhance IL-10 production [45]. Poly(cytidylic-inosinic) acid (poly(I:C)), a polyribonucleotide that mimics the effects of viral double-stranded RNA and activates TLR3, did not show any significant effect (Fig 6M).

**Fig 6.**
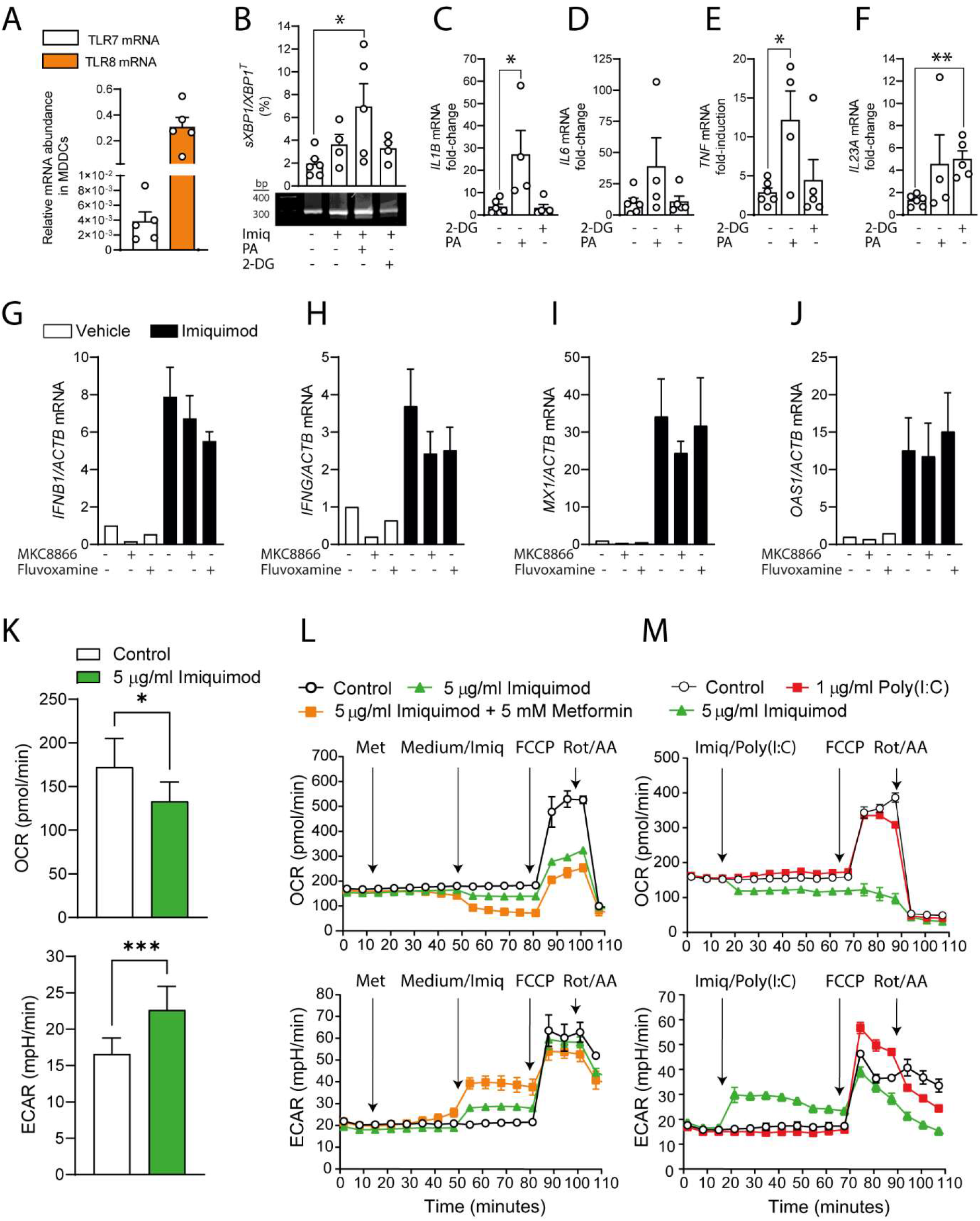
Expression of *TLR7* and *TLR8* receptors in MDDCs and response to the *TLR7* agonist imiquimod. (A) Expression of the mRNA encoding *TLR7* and *TLR8* in MDDCs. (B) Effect of 5 μg/ml imiquimod on *sXBP1* in the presence and absence of 0.5 mM palmitate (PA) and 10 mM 2-deoxyglucose (2-DG). MDDCs were incubated for one hour in the presence of the indicated additions and then stimulated with imiquimod for 1 hour. At the end of this time, the RNA was extracted and used for the assay of *sXBP1*(C-F) Effect of imiquimod on the expression of the mRNA encoding *IL1B*, *TNF*, *IL6*, and *IL23A*. The stimulation with imiquimod was maintained for four hours before RNA extraction. Data are presented as mean ± SEM. *p < 0.05, **p < 0.01. (G-J) Effect of MKC8866 and fluvoxamine on the expression of the mRNA encoding *IFNB1*, *IFNG*, *MX1*, and *OAS1*. MDDCs were preincubated with 10 μM MKC8866 and 20 μM fluvoxamine for one hour and then stimulated with 5 μg/ml imiquimod for 4 hours. At the end of this time, the RNA was extracted and used for mRNA assay. (K) Effect of imiquimod on O_2_ consumption rate (OCR) and glycolysis assayed as extracellular acidification rate (ECAR). Data are presented as mean ± SEM. *p < 0.05, ***p < 0.005, paired, (two-tail) Student’s t test. (L) Seahorse real-time metabolic analysis of one experiment where MDDCs were stimulated with imiquimod and then treated with 2 μM FCCP (an uncoupler of oxidative phosphorylation) to estimate maximal respiratory rate, and the combination 0.5 μM rotenone/antimycin (Rot/AA) to inhibit complex I and complex III of the mitochondrial electron transport chain. Where indicated, metformin was used to inhibit complex I activity. (M) MDDCs were treated with imiquimod or with poly(I:C) to show the effect of a TLR3 agonist.

In contrast to the limited effect of imiquimod, the *TLR8* agonist ssRNA40, a 20-mer phosphorothioate protected single-stranded RNA oligonucleotide containing a GU-rich sequence, induced *sXBP1* in a similar way to the effect of the TLR2 agonist zymosan. The splicing was blocked by the IRE1α RNase inhibitors MKC8866 and 4µ8C (Fig 7A). MKC8866 also inhibited the expression of *IL1B*, *IL6*, and *TNF* mRNA (Figs 7B-7D) and protein (Fig 7E-7F), thus resembling the cytokine signature detected in BAAs. ssRNA41, a ssRNA40 derivative wherein uracil nucleotides are replaced with adenosine and does not activate *TLR8*-dependent signaling, did not induce cytokine expression, induced *sXBP1* to a low extent, and was less active than ssRNA40 to induce the aggregation of misfolded proteins, as deemed from the assay of aggresomes (S1A and S1B Fig). While pro-IL-1β expression increased in response to ssRNA40 (Fig 7G), IL-1β was not detected in MDDCs supernatants, thus suggesting that ssRNA40 does not activate the inflammasome and that an additional signal(s) is required for IL-1β secretion. The expression of IL-6 and *TNF*α protein was inhibited by both MKC8866 and fluvoxamine, which suggests that *sXBP1* plays a significant role on the transcription of these cytokines. *IFNB1*, *MX1*, and *OAS1* mRNA were also induced by ssRNA40, while both MKC8866 and fluvoxamine only inhibited *MX1* and *OAS1* expression (Fig 7H-7K), thus suggesting a direct effect of sXBP1 on *MX1* and *OAS1* expression, rather than an indirect effect mediated by IFNs. Notably, ssRNA40 did not modify the energetic pattern of MDDCs (S1C Fig), which indicates that its ability to induce cytokine expression does not depend on blatant energetic changes. Unlike ssRNA40, zymosan, a ligand of TLR2 and C-type lectin receptors that mimics the external wall of fungi, induced a robust induction of glycolysis and OXPHOS as deemed from ECAR and OCR increases, respectively (S1D Fig).

**Fig 7.**
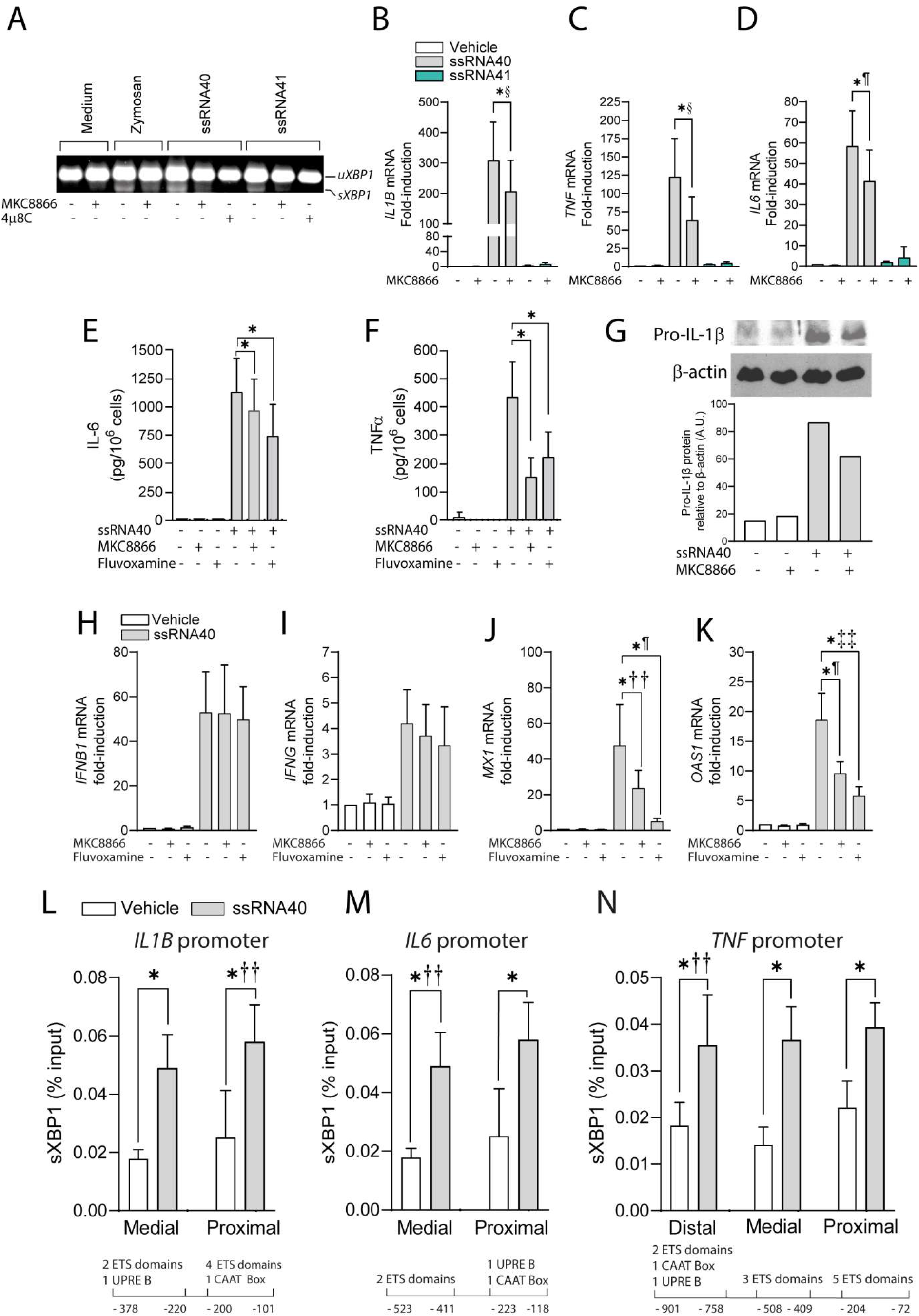
Effect of the *TLR8* agonist ssRNA40 on MDDCs. (A) Effect of ssRNA40 and IRE1α RNase inhibitors on *sXBP1*. The RNA was collected after one hour of incubation with 10 µM MKC8866 or 20 µM 4µ8C and one hour of stimulation with 2 μg/ml of either ssRNA40 or ssRNA41 and used for the assay of *uXBP1* and *sXBP1*(B-D) Effect of 10 μM MKC8866 on the mRNA expression of *IL1B*, *TNF*, and *IL6* mRNA. MDDCs were maintained for one hour in the presence of MKC8866 and then stimulated with ssRNA40 or ssRNA41 for 4 hours, prior to the extraction of the RNA for cytokine assays. Data are presented as mean ± SEM. *p < 0.05, **p < 0.01. §Wilcoxon matched pairs signed rank test. ¶Paired t test two-ways. (E and F) Effect of MKC8866 on the expression of IL-6 and *TNF*α protein. MDDCs were preincubated with MKC8866 for one hour and then stimulated overnight with ssRNA40. At the end of this period, supernatants were collected for cytokine ELISA assay. Data are presented as mean ± SEM. *p < 0.05. (G) Induction of the expression of pro-IL-1β by ssRNA40 and effect of MKC8866. MDDCs were treated as in the ELISA assays and after overnight incubation, cell extracts were collected and used for the assay of pro-IL-1β and β-actin proteins by Western blot. β-actin was used for normalization. A.U., arbitrary units. (H-K) Effect of MKC8866 and fluvoxamine on the expression of the mRNA encoding *IFNB1*, *IFNG*, *MX1* and *OAS1*. MDDCs were preincubated with 10 μM MKC8866 and 20 μM fluvoxamine for one hour and then stimulated with 2 μg/ml ssRNA40 for 4 hours. At the end of this time, the RNA was extracted for mRNA assay. Data are presented as mean ± SEM. *p < 0.05. ††Ratio paired t test. ¶Paired t test two-ways. ‡‡Mann Whitney test. (L-N) Effect of ssRNA40 on the binding of *sXBP1* to the promoters of *IL1B*, *IL6*, and *TNF*. The captions below the graphs indicate the distance from transcription start to the nucleotide positions where PCR primers were selected. The defined *sXBP1* binding sites included in the regions spanned by the primers are indicated. Samples were obtained after one hour stimulation by 2 μg/ml ssRNA40. Data are presented as mean ± SEM. *p < 0.05.††Ratio paired *t* test.

To confirm the involvement of sXBP1 in the transcriptional activation of cytokines, its binding to the proximal promoter regions of *IL1B*, *IL6*, and *TNF* was assayed. Chromatin immunoprecipitation (ChIP) assays were conducted with primers spanning areas including consensus *cis*-regulatory elements associated with position weight matrices discovered in sXBP1 target promoters [23]. An increased binding of *sXBP1* to the regions indicated in Fig 7L-N was observed after one hour of stimulation by ssRNA40. These results show that PAMPs acting on *TLR8* induce a cytokine signature like that observed in BAAs and point to the central involvement of MDDCs in the innate immune response to SARS-CoV-2. The presence of *sXBP1* in nasopharyngeal swabs and BAAs, its induction by ssRNA40 in MDDCs, the effect of IRE1α RNase inhibition on the cytokine induction produced by ssRNA40, and the demonstration of *sXBP1* binding to the *IL1B*, *IL6*, and *TNF* promoters suggest that *TLR8*-induced *XBP1* splicing may contribute to the viral sepsis observed in severe cases of COVID-19 disease.

## Discussion

Current pathogenetic views on COVID-19 pneumonia focus on immunopathological damage due to an exuberant innate immune response and a poor adaptive response. Assay of respiratory secretions allows the identification of the pathogens and can also give cues on pathogenesis [46–48]. This approach has been used in COVID-19 disease after the seminal studies by Zhou et al. [49] and Liao et al. [50], who used bronchoscopy and lavage to identify immune cell types in the respiratory tract. Our study focussed on patients under mechanical ventilatory support due to severe pneumonia, whose samples were obtained during routine care by attending staff [51–53]. This is in line with the use of tracheal aspirates to assess the transcriptional profiling of the lower respiratory tract in critically ill COVID-19 patients [54]. Initial assays showed higher degrees of *sXBP1* in COVID-19 disease than in patients with non-SARS-CoV-2 infection, although the mRNA levels of proinflammatory cytokines were higher in patients undergoing bacterial pneumonia. This was not fully unexpected, since cytokine storm is influenced by genetics and physiological conditions, in addition to cytokine levels [55, 56]. Moreover, COVID-19 patients received steroids in a regular schedule, which contributes to reduce proinflammatory cytokines, and showed an overall mortality lower than that observed in non-COVID patients. Notably, inhibition of IRE1α activity through activation of S1R by fluvoxamine protected mice from mortality during endotoxemia and fecal-induced peritonitis, as well as the production of IL-1β, IL-6, and IL-12 p40 by human leukocytes [14]. Consistent with the experimental results, early fluvoxamine treatment in individuals with mild COVID-19 illness was associated with a reduction of signs of clinical deterioration as compared to the placebo group [57]. Similar results were reported in the TOGETHER trial, which involved larger cohorts for study and showed a significant reduction of morbidity by fluvoxamine, as deemed from a reduced resort to either retention in a COVID-19 emergency setting or transfer to tertiary hospital. Consistent with these findings, an independent data safety monitoring committee recommended stopping randomly assigning patients to the fluvoxamine arm in view of the superiority criterion for the primary endpoint [58]. Stratification of patients showing active infection and *sXBP1* disclosed the association of *sXBP1* with higher levels of cytokine expression and their decrease after infection negativization, thus suggesting that the effect of fluvoxamine may be due to inhibition of the IRE1α-XBP1 branch.

Secondary goals of the study were the characterization of enzymes involved in the bioenergetics and the identification of myeloid-lineage differentiation footprints. Bioenergetic screening suggested active glycolysis during SARS-CoV-2 infection supported by HIF1 and elements of the malate-aspartate shuttle. However, the predominance of glycolytic enzymes cannot be straightforwardly construed as a proof of aerobic glycolysis or Warburg effect given the compromise of O_2_ supply associated with SARS-CoV-2 pneumonia. The strong induction of *IRG1*/ACOD1 mRNA is consistent with its dependence on IFNs [59, 60]. Notably, the ACOD1 product itaconate exerts antiviral activity and is considered a druggable target to counter the hyperinflammatory response [61].

Monocytic lineage cells are key players of the innate immune response due to their array of pattern recognition receptors and involvement in antigen presentation. BAAs showed a low expression of markers associated with antigen presentation, survival signals, and the ISG *MX1* during active infection. It is remarkable the low expression of *HLA-DRB1* mRNA, the gene encoding the most prevalent β-subunit of HLA-DR. This is in accordance with the decreased expression of HLA-DR in monocytes of COVID-19 patients, which drives hyperinflammation and defective antigen presentation mediated by IL-6 [62]. In contrast, the increased expression of *TMPRSS2* mRNA agrees with the facilitating role of this transmembrane protease in viral infection by cleaving viral S glycoprotein. Tellingly, single nucleotide polymorphisms at 21q22.3 locus within *TMPRSS2* and near *MX1* genes have been associated with severe COVID-19 disease [35].

*TLR7* and *TLR8* are tandem duplicated genes on the X-chromosome, the function of which shows some commonalities and specificities. For instance, *TLR8* is not functional in mice, and this explains the involvement of *TLR7*-induced cytokines in murine influenza [63]. *TLR8* expression is a hallmark of human MDDCs, while *TLR7* is present in monocytes, macrophages, and plasmacytoid dendritic cells [33]. A pioneering study addressing the pathophysiology of 2003 SARS-CoV-1 outbreak showed a unique ability of SARS-CoV-1 GU-rich RNA sequences to induce proinflammatory cytokines through *TLR7* in mice and *TLR8* in human leukocytes [64]. This notion was extended in a recent report by comparing the effect of GU-rich RNAs on inflammasome activation and proinflammatory cytokine production. Notably, GU-rich RNA from the SARS-CoV-2 spike protein triggered the greatest inflammatory in human macrophages via *TLR8* [65]. This agrees with our BAA and *in vitro* studies showing *TLR8* as a central element in the recognition of +ssRNA virus and suggests a unique involvement of MDDCs and TLR8 in *XBP1* splicing and hyperinflammation. Comparison of ssRNA40 and ssRNA41 effects show that *TLR8*-dependent signaling and *sXBP1* are critical for cytokine expression, given the lack of effect of ssRNA41.

Activation of *TLR7* by imiquimod induced a limited set of MDDC responses. However, it was remarkable the effect on energetic metabolism, characterized by a drop of the OCR and a parallel increase of ECAR, which mimicked the well-known effect of bacterial lipopolysaccharide. The effect on cytokines and *sXBP1* was negligible and only reached significant values in the presence of palmitate. Unfortunately, our study does not contribute to answer open questions regarding the actual role of *TLR7* in SARS-CoV-2 defense and immunopathology. *TLR7* mutations driving loss-of-function in the antiviral response have been associated with severe forms of COVID-19 disease in young male [28]. Another study showed that while some *TLR7* variants exhibit a robust loss-of-function on type I IFNs production, other variants only have a marginal effect, thus suggesting that *TLR7* may shape the anti-viral response through additional mechanisms [66]. Moreover, autosomic inborn errors of TLR3- and IRF7-dependent type I IFN immunity were found in 23 out of 659 patients with severe COVID-19 pneumonia [67], thus stressing the role of type I IFNs in the protection against severe forms of COVID-19 pneumonia. Our data agree with Ito et al. [68] findings, who first disclosed that the *TLR7*/*TLR8* agonist R848 was 100-fold as potent as imiquimod in human MDDCs. The absence of energetic rewiring induced by ssRNA40 could be explained by IFNβ effect, since IFNβ restrains aerobic glycolysis during mycobacteria infection. This drives mitochondrial stress and helps explain why type I IFN may cause damaging effects to the host [69]. Consistent with this interpretation is the lack of effect of poly(I:C), a selective activator of TLR3 that by using TRIF as the sole adaptor, activates IRF3 and ultimately induces type I IFNs. This is in shark contrast with the effect of zymosan, a ligand for TLR2 and the C-type lectin receptor dectin-1 [70]. The binding of *sXBP1* to *IL1B*, *IL6*, and *TNF* promoters induced by ssRNA40, together with the strong reduction of cytokine expression by an inhibitor of IRE1α RNase, further indicates that the IRE1α-*XBP1* branch underpins the production of cytokines via *TLR8*. These results assign to *TLR8* capacities previously reported for TLR2 and TLR4, where *sXBP1* is required for sustained production of proinflammatory cytokines. A corollary to these results is that inhibition of IRE1α RNase activity could be a therapeutic approach for severe COVID-19 disease.

## Materials and Methods

### Patients, Leukocyte Samples, and Ethic Statements

Nasopharyngeal samples were obtained from patients studied in different medical departments for symptoms consistent with SARS-CoV-2 infection at *Hospital Clínico Universitario de Valladolid*. In the case of patients with mechanical ventilation and intubation, samples were obtained by endotracheal aspirations to remove respiratory secretions as part of clinical care by the attending staff. This allows the obtention of material from the alveolar and respiratory bronchiole level. BAAs were directly transferred to the DNA/RNA extraction kit MagMAX^TM^ Pathogen RNA/DNA (Applied Biosystems) for the automated extraction machine Kingfisher Flex (Thermo Fisher Scientific). Infection diagnosis was obtained using a TaqPath^TM^ COVID-19 RT-PCR kit assay from Applied Biosystems that targets N, ORF1a, and S genes. Resolution of infection was confirmed by the analysis of samples collected four days after a positive test. BAAs from non-COVID-19-patients were obtained from samples collected for microbiological diagnosis in patients suffering from severe bacterial pneumonia and requiring ventilatory support and intubation at ICU. Lung protective ventilation of both COVID-19 and non-COVID-19 patients was performed according to the current guidelines on mechanical ventilation of acute respiratory distress syndrome in adult patients, which makes it unlike the induction of cytokine expression by mechanical ventilation [71]. The clinical part of the study was approved by the Ethics Committee of *Area de Salud Valladolid Este* (ref. PI-GR-20-2011 COVID). For *in vitro* experiments, MDDCs were obtained from human mononuclear cells collected from pooled buffy coats of healthy donors provided by *Centro de Hemoterapia y Hemodonación de Castilla y León Biobank*. The study was approved by the Bioethical Committee of the Spanish Council of Research (CSIC) and the written informed consent of all healthy donors was obtained at *Centro de Hemoterapia y Hemodonación de Castilla y León Biobank*. The researchers received the samples in an anonymous way. The process is documented by the Biobank authority according to the specific Spanish regulations. The ethics committee approved this procedure before starting the study. The differentiation of monocytes was carried out in the presence of GM-CSF and IL-4 for 5 days. Culture was carried out in RPMI 1640 medium containing 11.1 mM D-glucose and 4 mM L-glutamine. 10% FBS was maintained during the differentiation process and reduced to 2% at the start of experiments. Imiquimod (Sigma-Aldrich), ssRNA40/LyoVec™, and its negative control ssRNA41/LyoVec™ (InvivoGen) were used as *TLR7* and *TLR8* selective ligands in MDDCs. MKC8866 was from MedChemExpress.

### *XBP1* Splicing Assay

This was carried out by RT-PCRs using primers outside the spliced region. The PCR conditions were 5 min at 95°C (hot start), 45 cycles of denaturation at 95°C for 15 s, annealing at 60°C for 20 s and elongation at 72°C for 1 min. Final extension was carried out at 72°C for 5 min. Gel electrophoresis was carried out in 3% agarose and *sXBP1* and *uXBP1* bands were visualized by GelRed® staining and quantified using GelDoc Go Image System (Bio-Rad).

### Real-Time RT-PCR and Protein Assays

Total RNA obtained by automatic extraction was used for RT reactions. The resulting cDNA was amplified in a LightCycler® 480 equipment using SYBR Green I mix containing Hot Start polymerase. Cycling conditions were adapted to each set of primers. *ACTB* was used as a housekeeping gene to assess the relative abundance of the different mRNA using the comparative cycle threshold method. The procedure was used to assay *XBP1*, *DDIT3*, *IL1B*, *IL6*, *IL10*, *IL23A*, *TNF*, *SLC25A11*, *GLUT1*, *HK2*, *PFKB3*, *PDK4*, *IDH1*, *IDH2*, *SDHA*, *MDH2*, *HIF1A*, *TLR7*, *TLR8*, *HLA-DRB1*, *CD300E*, *CCR2*, *MMP9*, *BATF3*, *MX1*, *OAS1*, PTGS2, and *TMPRSS2* mRNA. Primer sequences are shown in Table 1IL-6 and *TNF*α proteins were assayed in supernatants of MDDCs stimulated with ssRNA40 using kits from Elabscience. Pro-IL-1B was assayed by Western blot using an antibody from Cell Signaling Technology.

**Table 1.**
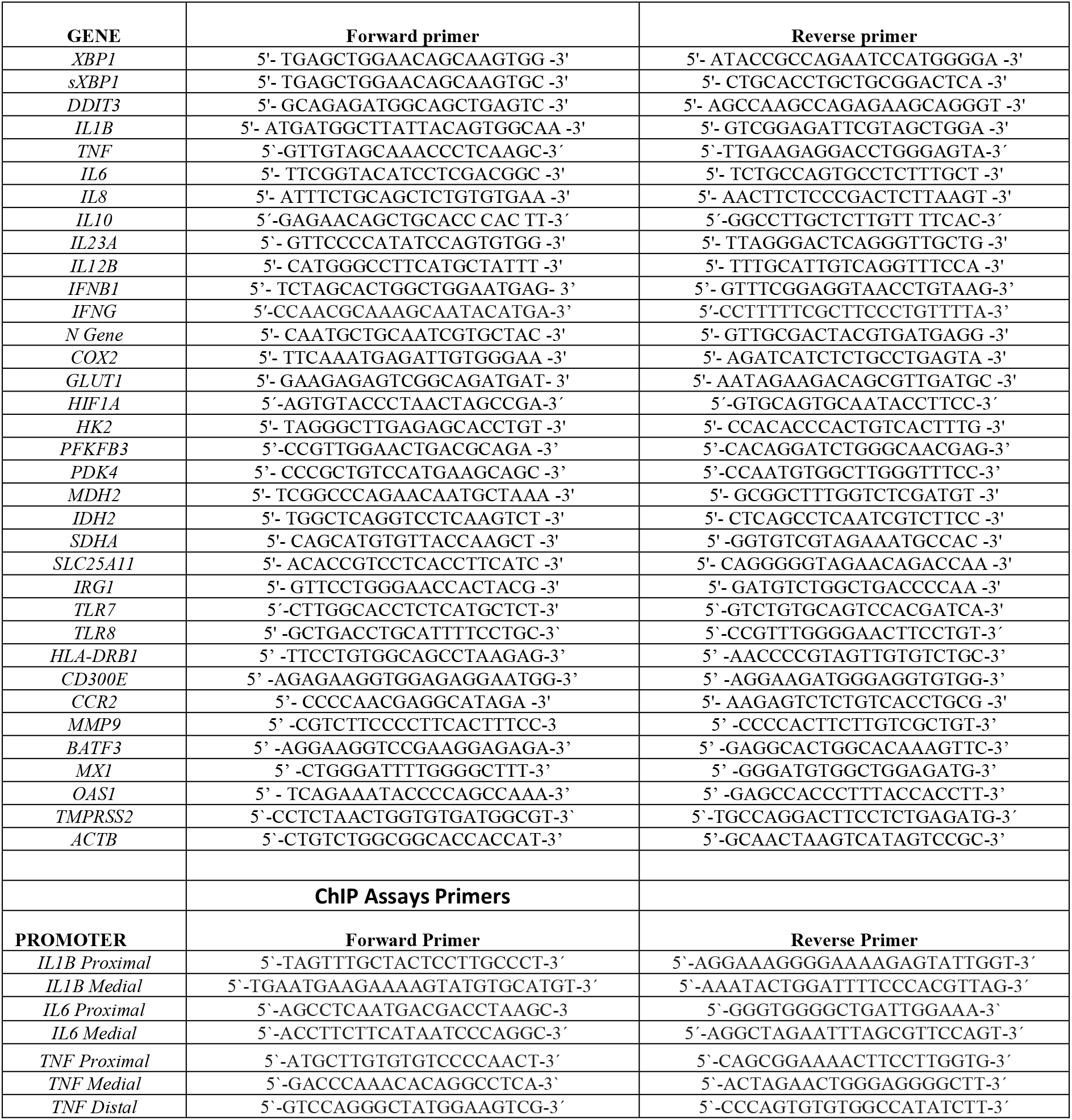
Sequences of primers used for qRT-PCR and ChIP assays.

### Real-Time Bioenergetic Analysis

Bioenergetic assays were carried out using an Agilent Seahorse XF HS Mini Analyzer. 10^5^ MDDCs were adhered with Cell-Tak® to Seahorse plates and treated with stimuli of TLR3, *TLR7*, and *TLR8*, as well as sonicated zymosan to activate the fungal pattern receptor dectin-1 and TLR2, after a stabilization period. OCR and ECAR were analyzed according to the XF Cell Mito Stress Test kit protocol in XF media under the experimental conditions and in response to metformin, oligomycin, FCCP, and rotenone plus antimycin A.

### Aggresome Formation Assay

The formation of aggresomes due to ER stress was assayed by flow cytometry fluorescence using the Proteostat® aggresome detection kit of ENZO according to the manufacturer’s instructions. The proteasome inhibitor MG132 was used as a positive control. MDDCs were incubated under different conditions and then fixed, stained with Proteostat® dye and used for the assay of fluorescence in a Gallios flow cytometer at 488 nm in the FL3 channel using the Kaluza software version 1.1 for quantitative analysis (Beckman Coulter Life Sciences).

### Chromatin Immunoprecipitation (ChIP) Assay

Chromatin immunoprecipitation assays were conducted using a rabbit mAb (Cell Signaling Technology) against *sXBP1* as previously reported [10]. Briefly, MDDCs were stimulated, washed with PBS, and fixed with 1% formaldehyde. Cross-linking was terminated by 0.125 M glycine. Crude nuclear extracts were collected by microcentrifugation. Chromatin sonication was carried out using a Bioruptor device (Diagenode). The chromatin solution was precleared by adding Protein A/G PLUS-Agarose for 30 min at 4°C under continuous rotation. After elimination of the beads, mAb was added for overnight incubation at 4°C, and then Protein A/G PLUS-Agarose was added and incubated for an additional period of 2 h at 4°C. Beads were harvested by centrifugation at 4,000 x g and sequentially washed with lysis buffer high salt, wash buffer, and elution buffer. Cross-links were reversed by heating at 67°C in a water bath, and the DNA bound to the beads isolated by extraction with phenol/chloroform/isoamylalcohol. Irrelevant Ab was used as control of binding specificity. The sequences of the primers are shown in Table 1Results are expressed as percentage of input.

### Quantification and Statistical Analysis

Data are represented as the mean ± SEM and were analyzed with the Prism 9.0 statistical program. Repeated measures one-way and two-way ANOVA analyses were performed. When data did not follow normal distribution nor had equal variances, log-transformation was applied before analysis. Comparison between experimental groups was carried out using unpaired or paired two-tailed Student’s t-test and Wilkoxon signed-rank test, and Welch’s test. Statistical details are shown in the Fig legends. Differences were considered significant for p < 0.05.

## Data Availability

All data produced in the present study are available upon reasonable request to the authors

## ACKNOWLEDGMENTS

José Javier Fernández is the recipient of a grant from *Junta de Castilla y León*. Cristina Mancebo is the recipient of a pre-doctoral grant from the Valladolid Section of Asociación *Española contra el Cáncer* (AECC). *Biobanco del Centro de Hemoterapia y Hemodonación de Castilla y León* is thanked for providing buffy coats. Staff from the Intensive Care Unit of *Hospital Clínico Universitario de Valladolid* is thanked for the effort devoted to patient follow-up care and sample collection. BioRender.com software was used in some figures.

## AUTHOR CONTRIBUTIONS

**Conceptualization:** Antonio Orduña, Juan Cubillos-Ruiz, Elena Bustamante, Nieves Fernández, Mariano Sánchez Crespo.

**Data curation:** José J. Fernández, Sonsoles Garcinuño, Gabriel March, Luis Inglada, Antonio Orduña, Juan Cubillos-Ruiz, Elena Bustamante, Mariano Sánchez Crespo.

**Formal analysis:** José J. Fernández, Yolanda Alvarez, Luis Inglada, Jesús Blanco, Antonio Orduña, Elena Bustamante, Nieves Fernández, Mariano Sánchez Crespo.

**Funding acquisition:** Nieves Fernández, Mariano Sánchez Crespo.

**Investigation:** José J. Fernández, Cristina Mancebo, Sonsoles Garcinuño, Gabriel March, Yolanda Alvarez, Sara Alonso, Luis Inglada, Jesús Blanco, Antonio Orduña, Olimpio Montero, Tito A. Sandoval, Juan Cubillos-Ruiz, Elena Bustamante, Nieves Fernández, Mariano Sánchez Crespo.

**Methodology:** José J. Fernández, Cristina Mancebo, Sonsoles Garcinuño, Gabriel March, Yolanda Alvarez, Sara Alonso, Olimpio Montero, Elena Bustamante, Luis Inglada, Jesús Blanco, Tito A. Sandoval, Antonio Orduña, Elena Bustamante.

**Project administration:** Nieves Fernández.

**Resources:** Nieves Fernández.

**Supervision:** Nieves Fernández, Mariano Sánchez Crespo.

**Validation:** Elena Bustamante, Luis Inglada, Jesús Blanco, Elena Bustamante, Nieves Fernández.

**Visualization:** José J. Fernández, Nieves Fernández, Mariano Sánchez Crespo.

**Writing – original draft:** Luis Inglada, Jesús Blanco, Tito A. Sandoval, Juan Cubillos-Ruiz, Nieves Fernández, Mariano Sánchez Crespo.

**Writing – review & editing:** Luis Inglada, Jesús Blanco, Tito A. Sandoval, Juan Cubillos-Ruiz, Elena Bustamante, Nieves Fernández, Mariano Sánchez Crespo.

