## Supplemental Figure 1 for "IRE1α-XBP1 Activation Elicited by Viral Singled Stranded RNA via *TLR8* May Modulate Lung Cytokine Induction in SARS-CoV-2 Pneumonia"

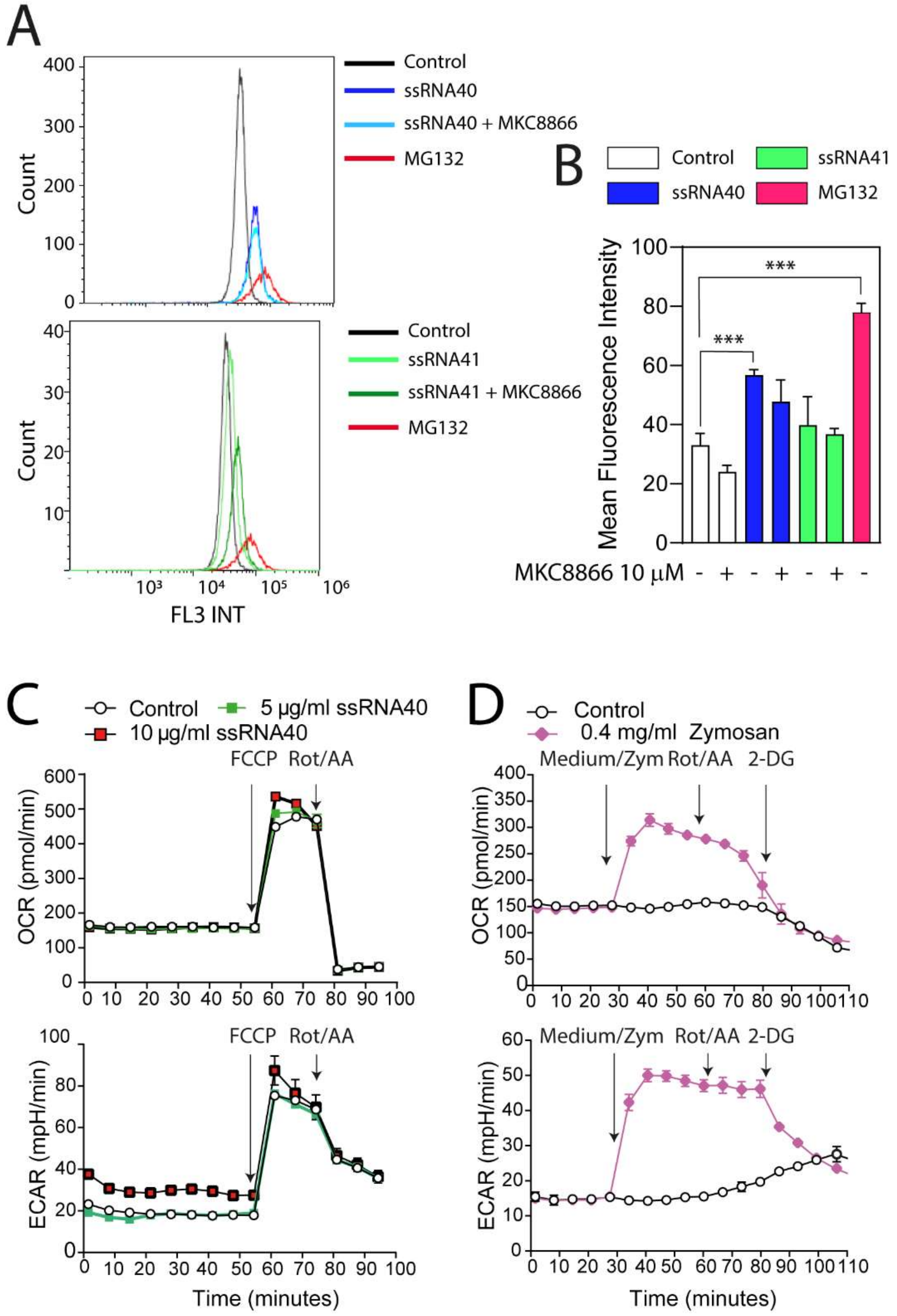

**S1 Fig. Effect of Effect of ssRNA40 on aggresome formation and energetic metabolism in MDDCs**. (A) Representative histograms of aggresome formation in MDDCs labeled with Proteostat® and stimulated for 4 hours with 2 μg/ml of ssRNA40 or ssRNA41 in the presence and absence of MKC8866, or with 5 μg/ml of the aggresome inducer MG132. After these treatments, MDDCs were fixed, stained, and analysed by flow cytometry using a 488 nm laser in the FL3 channel. (B) Mean fluorescence intensity of MDDCs stimulated overnight under the above-described conditions and stained with Proteostat®. Data are presented as mean ± SD. *** $p<0.005$, paired (two-tail) *t* test. (C) Effect of ssRNA40 on OCR and ECAR. MDDCs were stimulated with ssRNA40 and then treated with 2 μM FCCP to estimate maximal respiratory rate, and the combination 0.5 μM rotenone/antimycin (Rot/AA) followed by 2-deoxyglucose (2-DG) to assess proton exchange rate. (D) The fungal surrogate zymosan was used to show the robust activation of both glycolysis and OXPHOS induced through the activation of TLR2 and the C-type lectin receptor dectin-1.
